# Quality of Care for Children Under Five with Malaria Using IMCI Approach at Public Health Centre: Literature Review

**DOI:** 10.1101/2022.01.14.22269271

**Authors:** Orpa Diana Suek, Moses Glorino Rumambo Pandin

## Abstract

Integrated Management of Childhood Illness is one of the strategies in health care services for infants and children under five at primary health care facilities. Children with fever in high malaria endemic areas must have a blood test done to check whether the children have malaria or not. IMCI is one of the interventions recommended by WHO to screen and also ensure that children receive proper initial treatment at first-level health facilities. This article aims to discuss the quality of care for children under five with malaria using IMCI approach. This is a systematic review by searching 4 databases including Scopus, Web of Science, SAGE and Proquest. Health care services for children under five with an integrated management of childhood illness are intended to provide immediate and appropriate treatment. The guideline for treating children under five with malaria using IMCI approach is very helpful for nurses both in assessing, classifying, treating/curing and making decisions for pre-referral measures for severe cases. Several factors to support quality of care are trained officers, supervision and procurement of essential medicines, RDT and malaria microscopy. Enforcement of the right diagnosis will improve the quality of life of children and prevent death in children under five.

## INTRODUCTION

Various efforts are continuously made in order to reduce the mortality rate of newborns, infants and children under five. The health development goals stated in the Sustainable Development Goals (SDG’s) are to ensure a healthy life and promote well-being for all people at all ages, including infants and children under five who are prone to illness. One of the targets is ending infant and under-five mortality which can be prevented in 2030 by reducing the Neonatal Mortality Rate to at least 12 per 1000 Live Births and Children Under Five Mortality Rate to 25 per 1000 Live Births (Hussein, S., & Farhood, H. 2019; WHO, 2019).

Integrated Management of Childhood Illness (IMCI) has been introduced by WHO for 30 years ago with the aim at improving the health status of children. Currently, IMCI strategy is being performed all over the world including Indonesia. IMCI is an integrated strategy to improve children’s health, which is associated with a number of significant health problems as the main cause of morbidity and mortality such as pneumonia, diarrhoea, malnutrition, measles, malaria. Children brought to health care facilities are often found to be suffering from more than one unwell condition, making a single diagnosis impossible. Children need a combination therapy for successful treatment. Khan, MS, et al (2020) conducted a survey related to the implementation of IMCI and found that children require a combination therapy to achieve a successful treatment outcome.

Children under five with fever are screened with IMCI chart in a public health centre (PHC). Officers will carry out assessments, classifications, measures/treatments, counselling and decide on return visits (Ministry of Health, 2019; Suek, OD, & Ina, A. 2019). The patients with fever are screened for three diseases including malaria, measles and dengue hemorrhagic fever. In high malaria endemic areas, every sick child must be tested for malaria (Ministry of Health, 2021).

Malaria is an infectious disease caused by a parasite called Plasmodium that can infects humans through the bite of a female Anopheles mosquito that can attack anyone regardless of age, including children (WHO, 2019; Dittrich, S et al, 2020). Indonesia is a malaria endemic area, especially coastal areas. There are 5 (five) malaria endemic provinces in Indonesia. The implementation of IMCI needs to be developed. This intervention makes it easier for health workers to find cases of malaria in children and provide appropriate and rational treatment.

## OBJECTIVE

The objective of this paper is to explore quality of care for children under five with malaria using IMCI approach at public health centres covering assessment, classification and treatment.

## METHOD

The design of this study is a literature review. The literature used in this study consists of research articles related to quality of care for children under five with malaria using IMCI approach. These articles were published in reputable international journals. The literature was searched in Scopus, Web of Science, SAGE and Proquest databases. The keywords were IMCI, quality of care and malaria. The literature had to meet the inclusion criteria where the population should be children under five with fever or malaria. In addition, descriptive analysis was added.

**Table 1.1.**
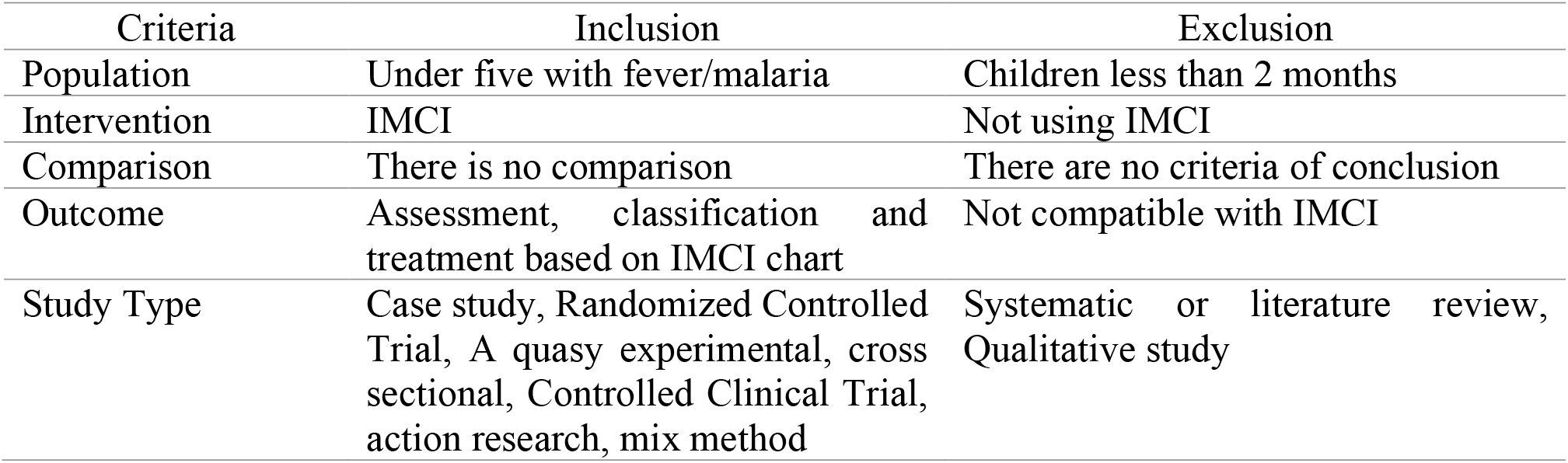
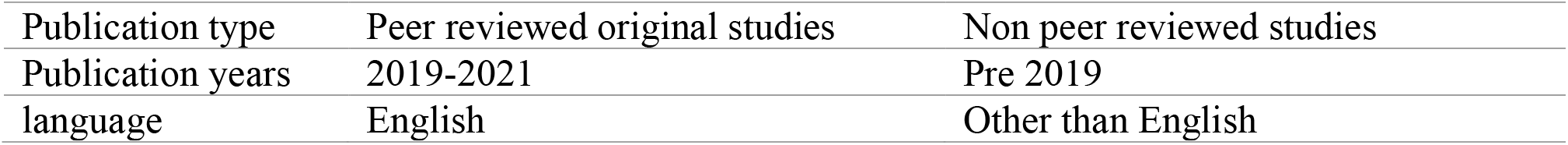
Inclusion and exclusion criteria with PICOS

**Figure 1.**
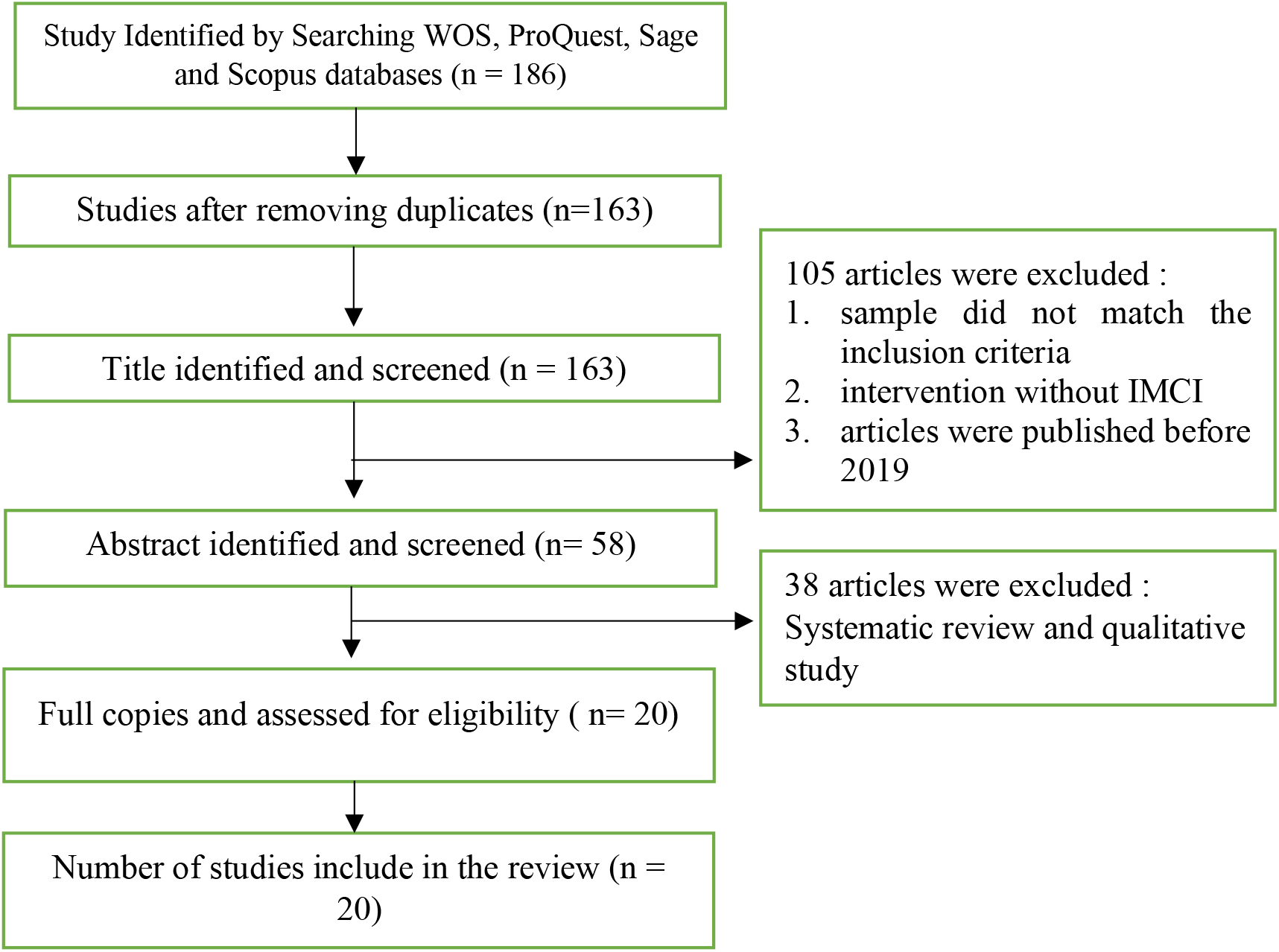
Article Selection Flow (PRISMA 2020)

## RESULTS

**Table 1.2.** Distribution of intervention using IMCI to improve quality of care for children under five with malaria.

| Researchers, Year | Result |
| --- | --- |
| Rozelle, JW et al (2021) | The presence of health assistants and the implementation of IMCI to children with malaria diagnosed 50% by confirmation of RDT and microscopic examination and 51% treatment of malaria in children under five in rural areas made the percentage of malaria diagnoses confirmed by microscopy or RDT increased from 71% before the program to 95% in 2020. |
| Abdulrahman, A., & Saleem, AM (2019) | The classification of sick children with IMCI guidelines has a significant effect due to a reduction of inappropriate use of antibiotics in children with fever |
| Nyaoke, BA, Mureithi, MW, & Beynon, C. (2019) | Prescription of antimalarial medicines despite negative microscopic results was found in 44 children (12.9%), with 48 mortality rate (14.1%) |
| Clarke-Deelder, E., Shapira, G., Samaha, H., Fritsche, GB, & Fink, G. (2019) | lack of adherence to diagnostic and treatment protocols. 332 (28%) of 1180 children under five who were observed, had signs of severe febrile disease, and less than 15% of patients were referred to a higher health care facility. |
| Getachew, T., Abebe, SM, Yitayal, M., Persson, L. ., & Berhanu, D. (2020) | Body temperature of patients (92%) were checked. It was found that 3% of children with fever were referred or treated, and 60% of them got antibiotics. 51% of children with malaria were assessed for screening with IMCI strategy and 4% of them were screened for general danger signs. |
| Hartley, MA, Hofmann, N., Keitel, K., Kagoro, F., Antunes Moniz, C., Mlaganile, T., & D'Acremont, V. (2020) | According to WHO Guidelines for Integrated Management of childhood Illness, it was recommended that antimalarial treatment was performed when the results of standard mRDT were released. Only a quarter of 2,801 outpatients with IMCI had parasites undetectable by standard mRDT. |

## DISCUSSION

The strategy for implementing IMCI includes promotive, preventive, curative and rehabilitative activities. IMCI services are focused on case management by looking at clinical signs and empirical therapy that can be carried out by doctors, nurses, midwives, and other health workers in primary services such as polyclinics, health centers, clinics, and hospitals. According to IMCI Handbook (Kemenkes, 2019), the process includes assessment and classification, which are interrelated and inseparable. IMCI must be performed comprehensively and simultaneously, especially in a developing country like Indonesia. This strategy strongly contributes to achieve children’s health status. Several articles claimed that IMCI was proven effectively to improve the quality of care for children, increase health care cost savings, and possibly reduce child mortality in developing countries.

Fever in children is one of the typical signs of infectious disease in children, where fever is said to be a general clinical sign of a disease contract. A study in malaria-endemic countries found that 71.4% of children with fever were caused by malaria Nkoka, O., Chuang, TW, & Chen, YH, 2019). In high malaria endemic areas, it is mandatory to check temperature and examine blood samples both by RDT and microscopic. IMCI is an evidence-based strategy that is widely adopted to prove testing and treatment for children with malaria, by improving access and quality of care specially to enforce diagnosis and treatment of malaria in children (Rozelle, et al. 2021). Balanza, N., et al, 2020 stated that “fever is a common problem in children. Most episodes of fever are caused by self-limiting infections, but a small number of children will develop life-threatening infections. Each infection has a symptom threshold, and other clinical symptoms can distinguish a sick child from asymptomatic or so-called progenic fever in malaria cases (Hartley, MA et al, 2020).

The protocol for a comprehensive and consistent integrated management of childhood illness (IMCI) has been used to promote accurate assessment and classification of childhood diseases, ensure appropriate combination treatment, strengthen caregiver counselling and accelerate referrals to reduce child mortality and morbidity. Success in implementing IMCI must be supported by training on IMCI standardization, weekly supportive supervision, availability of essential medicines and supporting infrastructure for IMCI services such as chart books and recording formulas.

In malaria endemic countries, malaria is suspected mainly by fever. WHO recommends early malaria diagnosis (within 24 hours of symptom onset) to prevent the occurrence of severe malaria. IMCI screening in febrile children has helped early diagnosis and treatment of uncomplicated malaria, thereby reducing the effects of severe malaria.

## CONCLUSION

IMCI is one of the appropriate interventions for screening, diagnostic tests and appropriate actions to children under five with malaria. Children are given antimalarial therapy if only supported by the results of RDT or malaria microscopy.

## Data Availability

All data produced in the present work are contained in the manuscript

